# Social Needs Z Codes and Subsequent Acute Care Across 21 U.S. Health Systems

**DOI:** 10.64898/2026.10.14.26365559

**Authors:** Felipe Lobelo, Jared Durieux, Sarmed Arshoo, Christos Colovos

## Abstract

Health systems increasingly record ICD-10 Z59 social-needs codes for housing, food, transportation, and related hardship to trigger referrals, report quality, and adjust risk. However code documentation occurs at varying rates and no study has separated what the code says about a patient’s hardship from what it says about being documented at all. Using a linked electronic health record and claims corpus covering 4.95 million patients in 21 identifiable health systems, we matched 26,153 adults with a first outpatient Z59 code to similar uncoded patients and, as a negative control, repeated the entire design around an administrative code. Coding ranged from 1 percent to 24 percent of patients across systems and remained highly variable after adjustment for patient mix. A hardship code was associated with 67% (95% CI=1.61, 1.74) more emergency department (ED) visits and 43% (95% CI=1.38, 1.49) more hospital admissions in the next year (62 versus 49 of every 100 patients with any acute event). The administrative-code reproduced 46 percent of the Z59 association for ED visits and 70 percent for admissions. In the five systems with the most complete stay dates, no hardship-specific admission excess was detectable. Homelessness codes forecast two and a half times the ED visit rate. At national planning prices, excess emergency and preventable hospital care totaled roughly $2,000–$2,500 per coded patient-year ($2,516 base scenario; about $1,100 under an exploratory assumed-discount scenario (not an estimate of hardship-attributable spending)), with the largest emergency burden among Medicaid patients. The Z code added no increment to clinical and utilization-based risk prediction models in any system.

## Introduction

The conditions in which people live (housing stability, food access, transportation, income) shape health and health care use.^1,2^ Health systems cannot treat those conditions directly in most encounters, but they can identify them, record them, refer patients to community resources, and account for them when they judge quality and risk. The National Academies recommended integrating social care into delivery,^3^ systematic reviews show a growing evidence base for screening and referral,^4^ and federal guidance now encourages recording social needs with ICD-10-CM Z codes, including the Z59 code family of housing and economic circumstances.^5,6^ Policy changes has, to some extent, increased the awareness and utilization of social needs coding as instrument that guides referrals, populates dashboards, feeds equity reports, and increasingly enters risk adjustment. Health systems and public programs are already acting on such flags at scale, including examples such as the Kaiser Permanente housing investments and medical-legal partnership work, food is medicine programming, county and state-based Housing for Health initiatives, and the Veterans Affairs Supportive Services for Veteran Families program, alongside navigation models such as the Accountable Health Communities model.^40–42^

For social and hardship codes to be documented four things have to coincide: (1) a patient has a need; (2) a clinician asks about it; (3) the workflow makes it easy to enter; and (4) the organization treats social-needs documentation as a priority. Coding therefore mixes the prevalence of hardship, the intensity of contact through which hardship can be noticed, and the documentation culture of the system. Every downstream use of hardship coding inherits that mixture. A system that codes 24 percent of its patients and a system that codes 1 percent are not describing the same flag, and a risk model or quality measure that treats the flag as interchangeable across systems will misread both.

Previous studies have established that patients with documented Z codes exhibit higher health care utilization. Bensken et al. found that inpatients with Z-code social needs in 2017 state discharge data had higher utilization and readmission;7 Liss and colleagues showed that any Z code in national commercial and Medicare Advantage claims predicted subsequent emergency and inpatient use, with socioeconomic codes carrying relatively higher inpatient risk.;^8,9^ Disease-cohort studies in older adults with heart failure and lung disease point the same way.^10^ Although social needs coding use has increased from 2015 through the early 2020s it still remains sparse,^11–13^ is highly variable by state, hospital, and system characteristics,^13–16^ and forecasts patients with more than twice the spending of uncoded patients.^12^ What previous studies have not established well is how much of the coded patient’s excess risk belongs to hardship and how much to the act of documentation itself, because to our knowledge no large-scale studies have compared estimates against a similar population with documented administrative Z codes not linked to social and hardship needs. Emergency physicians and other clinician and health system staff also record administrative Z codes, Z02 encounters for examination and Z76 encounters for other administrative purposes, through the same workflows, on the same mix of patients, with no plausible hardship pathway to subsequent health care utilization. Negative-control reasoning, a standard tool for pricing bias in observational studies,^17^ can therefore be used to compare the two associations; it does not by itself isolate documentation bias or identify a hardship-specific effect.

Under this context this study asks three policy questions the increasing use of social needs coding have made increasingly relevant. How much does coding vary across systems once patient mix is accounted for? Which specific hardships drive the risk of future healthcare utilization? And what is the ensuing acute care estimated cost, overall and by insurance, once documentation is discounted?. To address these questions we used a dataset derived from 21 U.S health systems in a linked electronic health record and claims corpus of 4.95 million patients.

## Methods

### Study Sample

This retrospective study used Protégé version 3 (snapshot June 4, 2026), a de-identified longitudinal database linking electronic health record (EHR) events to open medical and pharmacy claims and to a death file. The corpus derives from a single EHR aggregator contributing 39 health systems upstream; in the tables delivered for this analysis, 21 sites are identified. The delivery contains 4,950,000 patients residing in all 50 US states and the District of Columbia, enriched for diagnostic diversity by rare-code selection, so coding prevalences describe this corpus and not the US population. Structured data include demographics, encounters, diagnoses, procedures, laboratory results, vital signs, medication orders and fills, and surgical cases. Dates are shifted backward by a constant per-patient offset of 366 to 730 days, so within-patient intervals are exact but calendar time is approximate and for this reason, we do not report calendar dates of cohort entry or follow-up.

For this analysis the exposed cohort comprised adults aged 18 years or older whose first-ever social-needs documentation was a Z59 material-hardship code recorded on an outpatient or telehealth encounter, with no same-day ED or inpatient encounter, at least 365 days of prior observation, and no prior code from any social-needs code family. Comparators had no social-needs documentation of any family of codes through the end of their record and sought care via outpatient encounters in the same system and shifted year. A targeted sensitivity analysis repeated the comparison with comparator eligibility based only on information available through the index date (no listed family code on or before that date; later coding neither excluded the comparator nor triggered new censoring), preserving every other eligibility, matching, outcome, adjustment, censoring, and follow-up rule. Variable-ratio matching retained every exposed patient with at least one comparator, up to 1:4 without replacement. These comparators were matched on system and index year, a 5-year age band with a ±5-year caliper, sex, payer class at index, prior-year ED tier, admission tier, outpatient-volume tertile within system, and Elixhauser comorbidity tier.^18^ Of 27,683 eligible exposed patients, 26,153 (94.5%) retained at least one comparator.The primary analysis weights each comparator 1/k within its matched set, where k is the number of comparators in that set (98,869 comparator units; weighted total 26,153). Table 1 therefore describes 98,869 comparator units, not distinct comparator patients. Fixed 1:4 and 1:2 matching were used for sensitivity analyses. The negative-control cohort substitutes a first administrative code (Z02 or Z76, such as an encounter for examination or administrative purposes) for the hardship code and replicates every other analytic rule. Under this approach the analytic cohorts included 209,258 exposed patients and 688,392 comparator units. One data owner (rank 12) contributed coding-prevalence data but not matched cohorts because its records lacked the required linkage. Before delivery birth dates are reduced to the year, ages of 89 years and older are top-coded, residence is reported as state and 3-digit ZIP code, and names of people and places in clinical text are replaced with synthetic surrogates. An external review by Bluebonnet Ethical Review determined that this activity is appropriately classified as Not Human Subjects Research (NHSR) and that IRB review is not required.

**Table 1.**
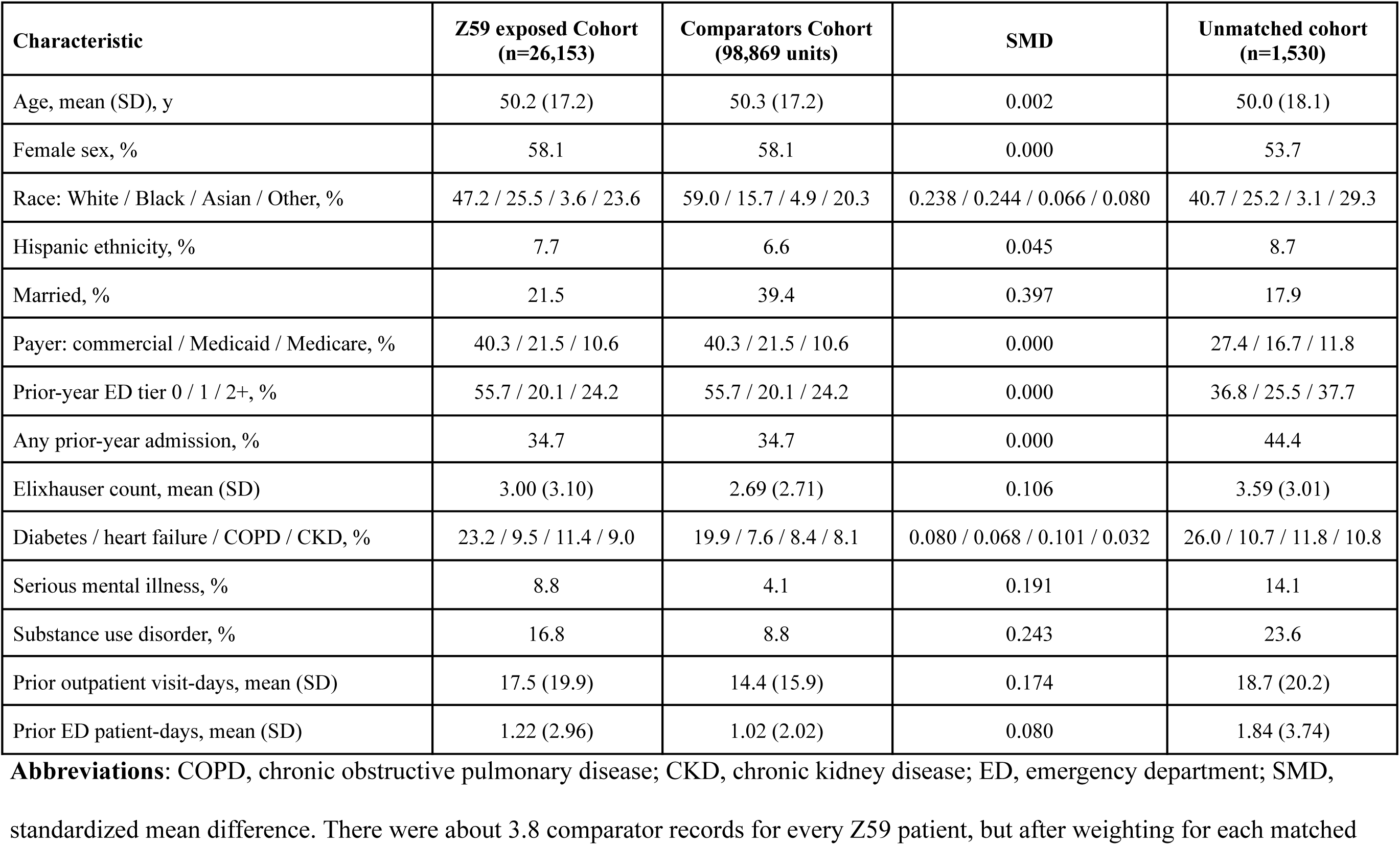
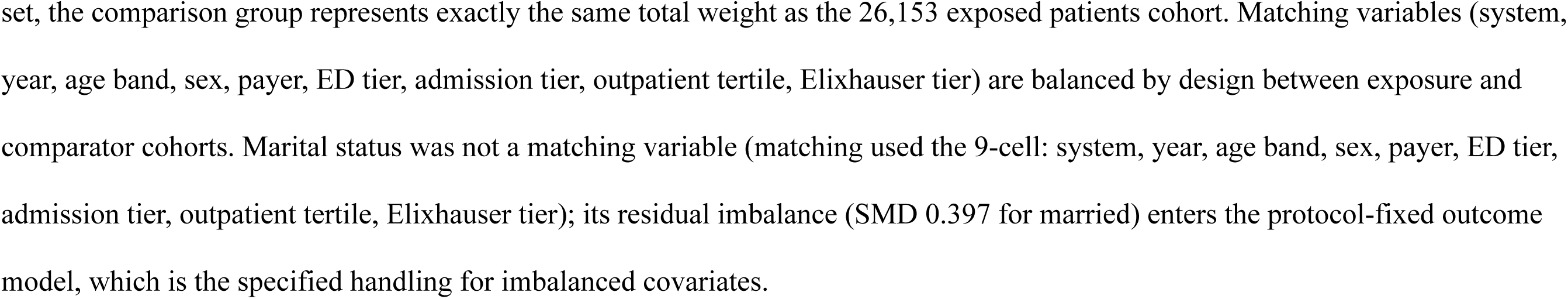
Baseline characteristics of matched patients, by social needs hardship-code status.

### Measures

The primary exposure is the first Z59 code (housing, food, transportation, financial, and related material-hardship codes; Appendix Table lists each code). Secondary exposures group the index code into housing, food, transportation, financial, and other subgroups, and a separate secondary exposure captures cost-related nonadherence (Z91.120, Z91.190). The negative-control exposure is the first Z02/Z76 administrative code.

Primary outcomes are counts of ED visit days and inpatient episodes during the 365 days after index encounter. The pipeline builds episodes from encounter records and claims through 2022, deduplicates events on patient-day, and counts an ED day inside a stay. Claims end in 2022 in each patient’s calendar date frame; after that date the record alone captures acute care, and an EHR-only sensitivity analysis tests that transition. A screened death endpoint requires a FACT_OF_DEATH record dated 2017 or later, corroboration by a second source within 31 days or a concordance with other coded structured data elements for death (e.g., claims, social security death verification), and no qualifying post-death activity more than 180 days later. Covariates come from the 12 months before the index encounter and include demographics, payer, marital status, area deprivation measures by ZIP3, comorbidities, serious mental illness, substance use disorder, and prior ED days, stays, and outpatient visits. The protocol fixed the outcome-model adjustment set before investigators saw any outcome. Covariates that remained imbalanced above a standardized mean difference of 0.10 were entered into a labeled sensitivity analysis only. Protocol-specified secondary outcomes are ambulatory-care-sensitive admissions under the Agency for Healthcare Research and Quality Prevention Quality Indicators, ICD-10-CM/PCS v2025 composite,^19^ and 30-day readmission. PQI v2025 specifications were verified code-for-code against the AHRQ release for PQI 05; Secondary outcomes use the merged episode rule supported by the admission audit (episodes beginning within 1 day of the previous episode’s end count as one stay).

### Cost Valuation

Modeled costs value measured excess events at external national prices. Excess ED visits and Prevention Quality Indicator admissions per 1,000 person-years (weighted crude, and assumed-discount counterparts applying the administrative-code comparison) were multiplied by planning prices in 2023 dollars: base $1,000 per ED visit and $15,000 per preventable stay, with low ($600; $11,000) and high ($1,600; $20,000) scenarios, and payer multipliers (Medicaid 0.80, commercial 1.25, Medicare and other 1.00) reflecting relative payment levels. Costs are gross associated excess over the 12 months after the index code encounter and not observed spending. Payer-stratified preventable-admission costs allocate each payer group’s excess admissions by the overall preventable share of excess admissions (42.5%) and are labeled modeled allocations.

### Statistical Analysis

Code-specific analyses repeat the primary rate models within each index-code group containing at least 1,000 exposed patients (homelessness, inadequate housing, food insecurity, transportation insecurity, financial strain), each against its own matched comparators; smaller code groupings were pooled as “Other”. These analyses were exploratory and hypothesis-generating. Rate ratios come from negative-binomial models with a log person-time offset, matched-set cluster-robust standard errors, and stabilized inverse-probability-of-censoring weights, constructed as described by Cole and Hernán,^20^ as part of the primary analysis (full-12-month observation initially differed between arms by more than 5 percentage points). An independent rerun corrected the weighting implementation. Primary intervals are from 500 matched-set bootstrap resamples refitting the censoring models; a consistent event/person-time Poisson analysis is reported as a sensitivity. The appendix details the censoring model, weight capping, and bootstrap. Cumulative incidence uses an Aalen–Johansen estimator^21^ with death as the competing event, a common observation endpoint for acute events and death, and 500 matched-set bootstrap resamples for the risk-difference interval. E-values follow VanderWeele and Ding.^22^

Between-system variation uses a multilevel logistic model of any first social-needs code with patients nested in systems. The model reports the intraclass correlation and the median OR with and without patient mix (age, sex, race, ethnicity, payer, comorbidity count, and prior utilization), and it compares system rankings before and after adjustment.^23^ The prediction analysis uses the unmatched eligible population with one index per patient per shifted year (the first outpatient encounter of the year). A baseline logistic model uses the protocol-specified covariates (demographics, payer, marital status, area measures, comorbidity, serious mental illness, substance use disorder, and prior utilization); an extended model adds a prior-12-month Z59 indicator. The analysis collapses demographic categories under 1% into Other or Unknown so that leave-one-system-out validation runs, and it reports ΔAUC, integrated discrimination improvement, net reclassification at 5%, 10%, and 20% risk thresholds,^24^ calibration, a decision curve,^25^ and leave-one-system-out ΔAUC by system. The earlier matched-cohort analysis remains as a sensitivity analysis. Prediction reporting follows the TRIPOD principles for model validation.^28^ Analyses used R version 4.1.2 (R Foundation for Statistical Computing, Vienna, Austria) and Presto SQL on the Protégé snapshot. Reporting follows the STROBE statement for cohort studies^26^ and the RECORD extension^27^; the completed STROBE checklist accompanies submission.

## Results

Coding prevalence ranged from 1.0% to 23.8% of adult patients across the 21 health systems. In the multilevel model, the between-system variance was 0.566, an intraclass correlation of 0.147 and a median OR of 2.05. Adjustment for patient mix reduced the variance to 0.397 (intraclass correlation 0.108; median OR 1.82), so case mix explained about 30% of the between-system variance. Crude and adjusted system rankings correlate at 0.90; the largest falls after adjustment are rank 15 (9th to 15th) and rank 2 (6th to 9th), and rank 18 remains the highest-coding system. Figure 1 displays crude prevalence by system. In the five systems with the most complete stay dates, no hardship-specific admission excess was detectable (Supplement, Appendix Figure 1). System coding tertiles, fixed before outcome analysis from these prevalences, structure the heterogeneity analysis below.

**Figure 1.**
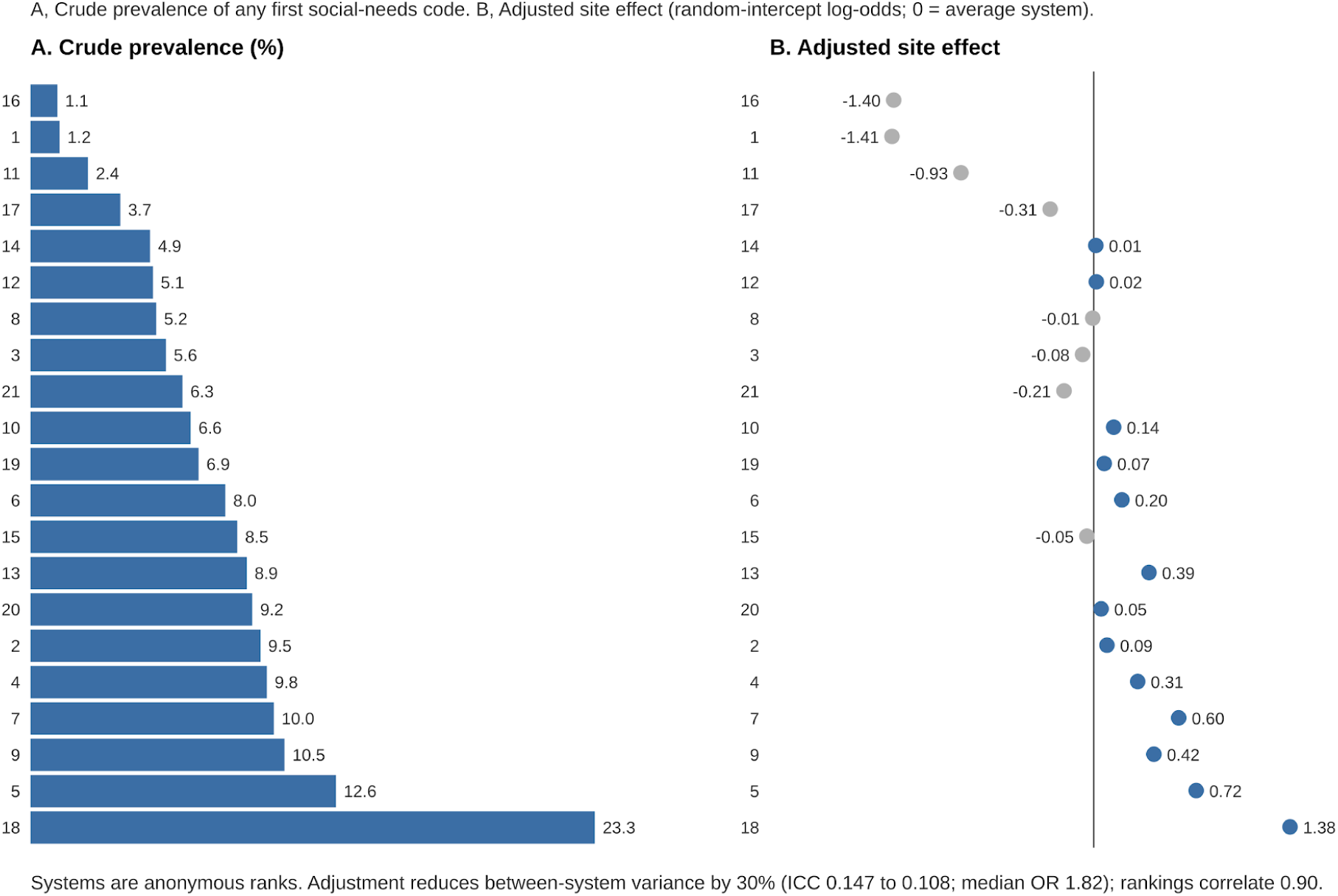
Coding prevalence and adjusted system effects. Panel A shows the crude percentage of adult patients with any first social-needs code in each of the 21 systems. Panel B shows patient-mix-adjusted system random effects on the log-odds scale from a multilevel logistic model adjusting for age, sex, race, ethnicity, payer, comorbidity count, and prior utilization; zero is the average system, so bars to the right identify systems that code more than their patient mix predicts. The between-system intraclass correlation falls from 0.15 unadjusted to 0.11 adjusted (median odds ratio 1.82), and crude and adjusted rankings correlate at 0.90. Systems are anonymized by rank

### Acute care after a hardship code

Patients in the exposed cohort generated 1,294 ED visits and 1,333 admissions per 1,000 person-years, compared with 640 and 1,041 among the comparator cohort. The adjusted rate ratio was 1.67 (bootstrap 95% CI=1.61, 1.74) for ED visits and 1.43 (bootstrap 95% CI=1.38, 1.49) for admissions (Table 2). A consistent event/person-time (Poisson) sensitivity analysis, which changes the model as well as the weights, gives 1.60 (95% CI=1.54, 1.75) and 1.26 (95% CI=1.18, 1.35). The admission episode audit found that 53% of exposed and 46% of comparator EHR episodes were built from valid stay dates, that admitted patients averaged 3.3 admission episodes in the year in both arms, and that only about 1% of episodes began within 1 day of the previous episode’s end. Merging such episodes lowers admission rates by about 5% in both arms (from 1,399 to 1,333 and from 1,097 to 1,041 per 1,000 person-years) and leaves the rate ratio at 1.41, against 1.42 under the original rule. In the 5 systems where at least half of inpatient rows carry valid stay dates, admission rates are higher in both arms and the adjusted admission rate ratio is 1.16 (95% CI=1.08, 1.24), while the negative control in the same systems is 1.29 (95% CI=1.26, 1.32); within those systems the hardship-specific admission excess was therefore not detectable (Supplement, Appendix Figure 1).

**Table 2.** Twelve-month outcomes and the administrative-code comparison.

| **Outcome** | **Z59 hardship code cohort, RR (95% CI)** | **Z02/Z76 comparison cohort, RR (95% CI)** | **Ratio comparing the two associations (95% CI)** | **Observed-comparison share of Z59 log-association** |
| --- | --- | --- | --- | --- |
| ED visits | 1.67 (1.61, 1.74) | 1.26 (1.25, 1.28) | 1.32 (1.27, 1.38) | 46% |
| Admissions, merged episodes | 1.43 (1.38, 1.49) | 1.29 (1.27, 1.30) | 1.11 (1.07, 1.16) | 70% |
| PQI composite admissions | 1.68 (1.59, 1.78) | 1.38 (1.35, 1.41) | 1.22 (1.15, 1.30) | 61% |
| PQI 10 sensitivity | 1.78 (1.62, 1.96) | 1.42 (1.35, 1.49) | 1.26 (1.13, 1.40) | 61% |
| 30-day readmission episodes | 1.50 (1.39, 1.62) | 1.43 (1.40, 1.46) | 1.05 (0.97, 1.14) | 87% |
| Any readmission among admitted (OR) | 1.20 (1.17, 1.24) | 1.26 (1.25, 1.27) | 0.96 (0.93, 0.99) | >100% |

**Abbreviations:** CI, confidence interval; ED, emergency department; OR, odds ratio; PQI, Prevention Quality Indicator. Ratio CIs treat the two cohorts as independent and derive standard errors from the reported intervals; cross-cohort patient overlap could not be verified from the deposited record, so that independence assumption is unverified. The administrative-code comparison is the share of the Z59 log-association numerically reproduced by an administrative code estimated under identical rules (an observed-comparison share, not an identified documentation effect), and the ratio comparing the two associations is Z59 exposed cohort over comparison cohort.

The 12-month cumulative incidence of a first acute event reached 62.4% in exposed patients and 49.1% in comparators in the corrected Aalen-Johansen analysis, a risk difference of 13.3 percentage points (95% CI=12.6, 13.9). Death before any acute event remained uncommon (0.3% versus 0.5%), and screened death within 12 months was nearly identical between arms (1.54% versus 1.56%). Homelessness codes carried the strongest association (ED rate ratio 2.49; admissions 1.86; Figure 3), followed by inadequate housing, food, transportation, financial, and other Z59 groups. Admission rate ratios rose across coding tertiles, from 1.20 in low-coding systems to 1.36 in middle-tertile systems and 1.48 in high-coding systems (interaction P=0.0006; Figure 2). ED rate ratios did not vary by tertile (interaction P=0.24).

**Figure 2.**
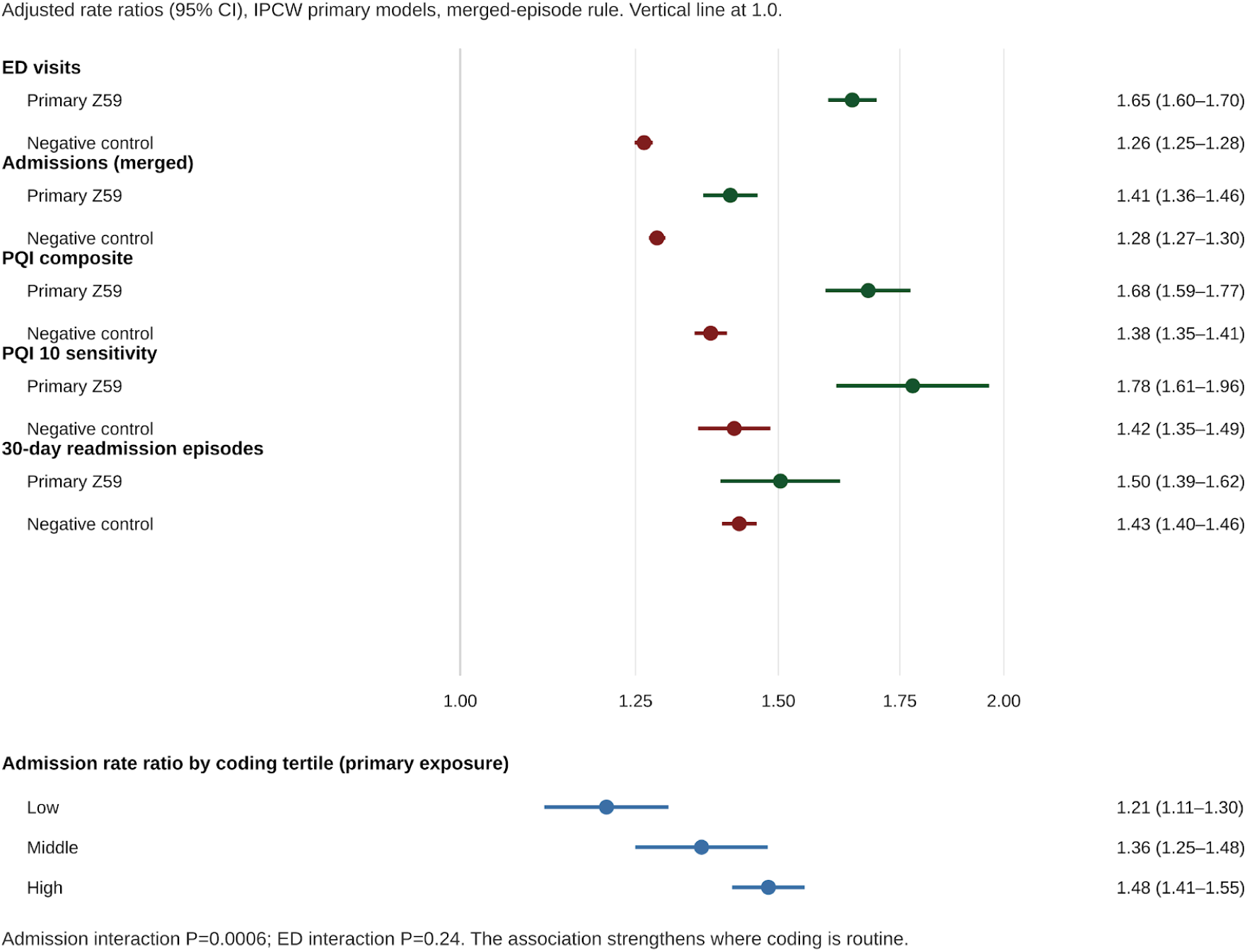
Hardship codes versus paperwork codes, and the coding-intensity gradient. Panel A (forest plot) shows adjusted rate ratios, each from an identical matched-cohort model with inverse-probability-of-censoring weighting, for a first Z59 hardship code (blue) and a first Z02/Z76 administrative code (grey, the negative control) across emergency department visits, admissions under the merged-episode rule, Prevention Quality Indicator composite admissions, the PQI 10 sensitivity, and 30-day readmission episodes. Panel B shows the primary admission rate ratio rising across low, middle, and high system coding tertiles (1.20, 1.36, 1.48; interaction P=0.0006)

**Figure 3.**
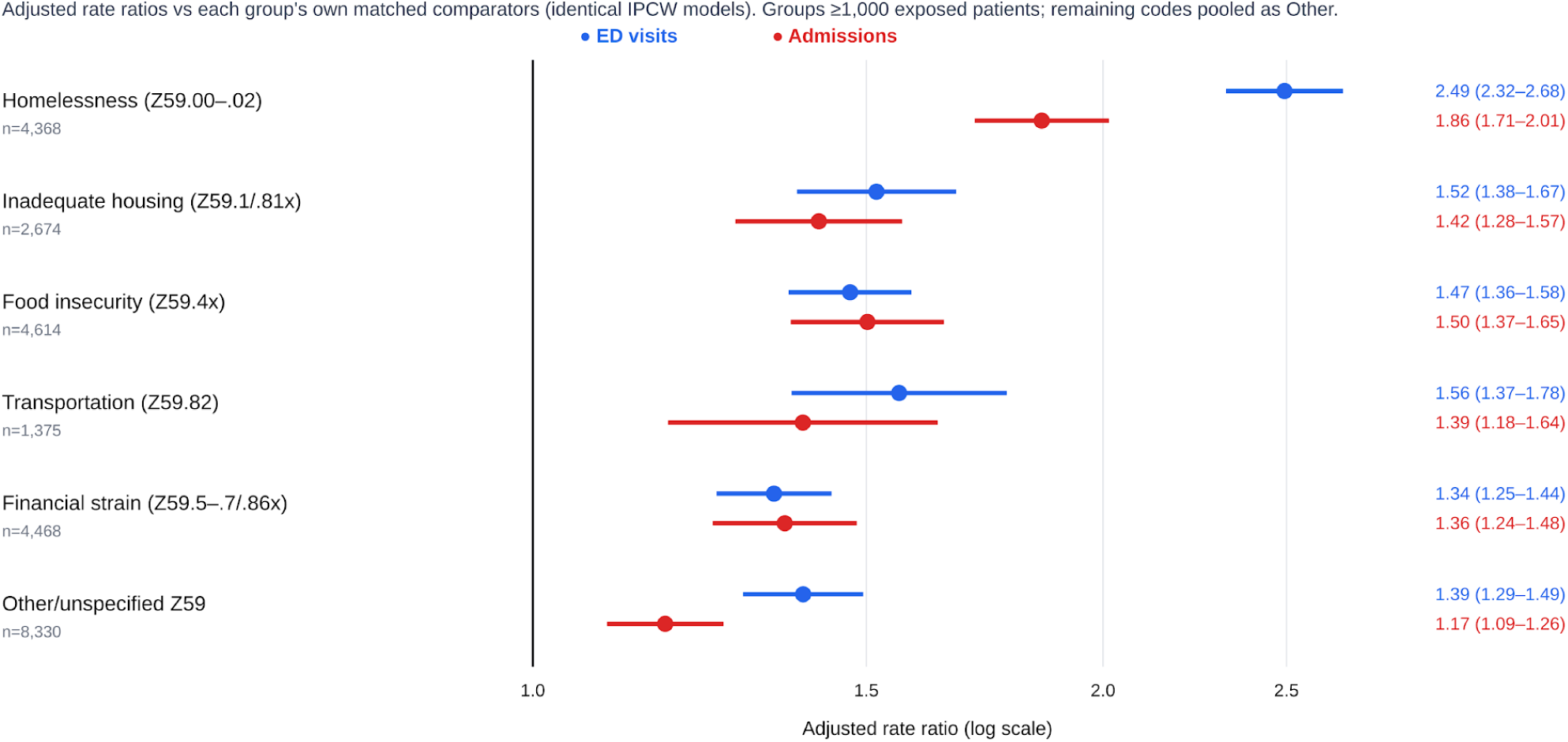
Which hardship was recorded. Adjusted emergency department (blue) and admission (red) rate ratios for each index-code group containing at least 1,000 exposed patients, each against its own matched comparators under the primary model. Homelessness codes (Z59.00 to Z59.02) carry the strongest signal (2.49 and 1.86); food insecurity is the only group whose admission ratio meets its emergency ratio. Groups below 1,000 patients were pooled as Other to avoid unstable, multiplicity-prone estimates. Index-code group counts (4,436 + 2,709 + 4,666 + 1,394 + 4,544 + 8,404 = 26,153) are pre-model assignments; modeled matched sets sum to 26,150 and analyzable rows to 25,829 after requiring positive person-days (3 sets / 324 rows without observable follow-up). No estimate uses the pre-model counts.

### The negative control

A first administrative code also predicted acute care, at rate ratios of 1.26 (95% CI=1.25, 1.28) for ED visits and 1.29 (95% CI=1.28, 1.31) for admissions, with a first-event risk difference of 6.1 percentage points (95% CI=5.9, 6.3). The administrative-code association numerically reproduced 46% of the Z59 log-association for ED visits and 70% for admissions. These are observed-comparison shares, not identified fractions of excess events attributable to documentation. The ratio comparing the two associations is 1.32 for ED visits and 1.11 for admissions. Administrative-code cohorts also had higher subsequent acute-care use. The ratios comparing the two associations provide a sensitivity comparison; they do not isolate documentation bias or identify an effect attributable to hardship. Discounted costs are assumption-based scenarios, not estimates of hardship-attributable spending. Under the merged analytic rule, the negative-control admission rate ratio is 1.29 (95% CI=1.27, 1.30), unchanged, and in the valid-date systems it is 1.29 (95% CI=1.26, 1.32), above the primary contrast of 1.16 in those systems. In the targeted index-time sensitivity (26,694 exposed patients retained; 101,546 comparator units), re-estimated with the same deposited pipeline, the Z59 associations were attenuated but persisted: ED visit rate ratio 1.55 (95% CI=1.50, 1.60) and admission rate ratio 1.33 (95% CI=1.28, 1.37), against 1.65 and 1.42 under the original through-end-of-record eligibility in the same pipeline, with a first-event risk difference of 11.7 percentage points (95% CI=11.0, 12.3) against 13.1 points. Restricting comparator eligibility to information available at index therefore shrinks, but does not remove, the association (Supplement Table S3).

### Sensitivity analyses and prediction increment

Stratified sensitivities (labeled, post-protocol; Supplement Table S5) did not overturn the primary contrasts. By payer, associations were strongest in Medicaid (ED rate ratio 1.71, 95% CI=1.61, 1.81; admissions 1.48 (95% CI=1.38, 1.59)), intermediate in commercial insurance coverage (1.64; 1.35) and Medicare (1.45; 1.27), and high in the heterogeneous other/missing insurance product line group (1.70; 1.62). In analyses using ZIP3 data, low-deprivation areas showed the strongest associations (ED 1.81, 1.71, 1.91; admissions 1.56, 1.45, 1.69), followed by mid (1.65; 1.39) and high deprivation (1.45; 1.34). That inversion is consistent with an individual code marking more selected hardship where area deprivation is low, and with ZIP3 being too coarse to price individual need. Estimates were stable by era (2013–2019: ED 1.69, admissions 1.37; 2020–2026: 1.63, 1.43).

In the protocol population (298,070 patient-years; 1,602 with a prior-12-month Z59 code), the baseline model reached an area under the curve (AUC) of 0.7594. Adding the Z59 indicator did not change the AUC estimate. In the matched-cohort sensitivity analysis, the baseline AUC was 0.7414 and adding the Z code also did not contribute predictive value.

### Secondary outcomes and sensitivity analysis

Ambulatory-care-sensitive (Prevention Quality Indicator; PQI v2025 composite) admissions occurred at 315 versus 191 per 1,000 person-years after a Z59 hardship code, an adjusted rate ratio of 1.68 (95% CI=1.59, 1.78). Thirty-day readmission episodes occurred at 605 versus 481 per 1,000 person-years (rate ratio 1.50, 95% CI=1.39, 1.62), and among admitted patients the odds ratio for any 30-day readmission was 1.20 (95% CI=1.17, 1.24). After an administrative Z code, the corresponding rate ratios were 1.38 (PQI composite), 1.42 (PQI 10), and 1.43 (readmission episodes), with an among-admitted odds ratio of 1.26. Preventable admissions therefore separated hardship from documentation somewhat better than all-cause admissions (ratio of rate ratios 1.22 versus 1.11), and readmission did not separate them (1.05).

Unweighted estimates matched the weighted primary analysis (ED 1.66; admissions 1.42). EHR-only outcomes estimates were 1.70 and 1.41. Complete 1:4 matched sets estimates were 1.68 and 1.42 while analyses of 1:2 matched sets were 1.66 and 1.41. Exclusion of the 2 lowest-retention systems (ranks 18 and 21) resulted in comparable (1.66 and 1.42) estimates. Adding 5 further imbalanced covariates outside the fixed adjustment set attenuated estimates slightly (1.61 and 1.41).

Homelessness codes (Z59.00–Z59.02; 4,436 patients) carried by far the strongest signal: an adjusted ED rate ratio of 2.49 (95% CI=2.32, 2.68) and an admission rate ratio of 1.86 (1.71, 2.01) against their own matched comparators. Inadequate-housing codes followed (1.52; 1.42), then food insecurity (1.47; 1.50), transportation insecurity (1.56; 1.39), financial strain (1.34; 1.36), and other or unspecified codes (1.39; 1.17). Every code grouping included at least 1,300 patients and individual codes below that threshold were pooled. (Figure 3).

Valued at national planning prices, the excess acute care in the year after a hardship code totals about $2,516 per coded patient (base prices: $654 for ED visits plus $1,862 for preventable admissions), ranging from $1,758 at low prices to $3,529 at high prices (Table 3). Applying the assumed discount gives about $1,074 per patient-year (range $739–$1,525) as an exploratory assumed-discount scenario, not an estimate of hardship-attributable spending. By insurance, the ED component was largest among Medicaid patients ($763 gross per patient-year at payer-adjusted prices; excess 954 ED visits per 1,000 person-years) and the preventable-admission burden, allocated by payer, remained the dominant driver of cost in every group. All-cause admission valuation roughly doubled the totals and is shown only as an upper bound. These are gross associated amounts in an enriched, high-utilizing matched population, a planning figure for the scale of post-code acute care, not an estimate of savings from coding, screening, or referral.

**Table 3.** Modeled excess costs in the 12 months after a first Z59 code.

| **Component** | **Excess events**<br>**/1,000 py** | **Gross costs per**<br>**coded patient-year** | **Assumed-discount**<br>**scenario costs**<br>**(exploratory; not**<br>**hardship-attribut**<br>**able estimates)** |
| --- | --- | --- | --- |
| Emergency department visits | 654 | \$654 | \$355 |
| Preventable admissions | 124 | \$1,862 | \$719 |
| Total, base prices (\$1,000 ED; \$15,000 stay) | — | \$2,516 | \$1,074 |
| Total, low prices (\$600; \$11,000) | — | \$1,758 | \$739 |
| Total, high prices (\$1,600; \$20,000) | — | \$3,529 | \$1,525 |

## Discussion

In this analysis of EHR and claims data from 21 health systems, social risk coding prevalence ranged from 1.0% to 23.8% of adult patients, and patient mix explained only about 30% of that between-system variance. An initial Z59 code documentation was a common, consequential, and unevenly applied risk flag. About 62 of every 100 patients with Z codes had an ED visit or an admission within a year, 13 more per 100 than matched comparators. An administrative Z code, which should carry no hardship content, reproduced about half of the ED and about three quarters of the admission log-association. Preventable admissions separated the two exposures somewhat better than all-cause admissions did but readmission did not separate them at all. In the full eligible population, a prior Z59 code added no measurable discrimination to a model that already included age, comorbidity, payer, and prior utilization.

A common interpretive problem in social risk coding research is that the exposed group is defined by an act of documentation. Patients who receive a Z59 code have, by default, had an encounter in which social risk was elicited, recognized, and recorded. Those patients also tend to have more encounters, more diagnoses per encounter, and more prior acute care, known factors that predict future acute care. Conventional adjustment partially accounts for the measured component of that problem. Matching in this study balanced system, year, age band, sex, payer, comorbidity tier, and prior-use tiers, and the outcome model adjusted for the protocol-fixed set of marital status, race, Hispanic ethnicity, substance use disorder, serious mental illness, and prior ED days, stays, and outpatient visits. What adjustments are usually not fully accounting for is the residual tendency of some patients – and some clinicians and systems – to generate more documentation per unit of encounter. The negative control attempts to quantify that residual tendency. Because the administrative-code cohort was built with the same outpatient index rule, the same lookback, the same matching cells, the same outcome construction, and the same models, its rate ratios of 1.26 for ED visits and 1.29 for admissions provide a sensitivity comparison; they do not isolate documentation bias or identify an effect attributable to hardship. The social hardship codes outpredict it clearly for future ED utilization (ratio of rate ratios 1.32) and narrowly for admissions (1.11). Therefore two conclusions can be inferred from this comparison. First, the Z59 association is not an artifact of documentation alone; if it were, the negative control would have reproduced it at full size. Second, admission associations reported without the administrative-code comparison should not be read as hardship-specific amounts; the ratio comparing the two associations is 1.11 for admissions. That is a stronger and more specific insight than traditionally mentioned limitations about residual confounding, because it quantifies the bias of greatest concern.^17^

The same logic should be used when interpreting secondary outcomes. We hypothesized that ambulatory-care-sensitive admissions would separate hardship from documentation better than all-cause admissions, on the grounds that housing instability, food insecurity, and transportation failure should plausibly convert manageable illness into preventable hospitalization more than they convert it into elective admission. The data support that hypothesis only modestly: the ratio of rate ratios is 1.22 for the Prevention Quality Indicator composite (1.68 versus 1.38) and 1.11 for all-cause admissions. Thirty-day readmission, by contrast, does not separate the exposures (episode rate ratios 1.50 versus 1.43; among admitted patients, odds ratios 1.20 versus 1.26).

Readmission is heavily driven by the index admission, discharge processes, and the intensity with which a system documents and follows its admitted patients – all features shared by the two exposures. Preventable admission retains a hardship gradient, but even there an administrative-only code already captures most of the risk. Quality programs that treat coded preventable admissions as a pure measure of unmet social need will therefore misread a documentation signal as a needs signal in at least some of their patients.

The 23-fold range in crude coding prevalence is the most important descriptive estimate in this study, and the multilevel model shows that it is not mainly due to case-mix. Adjustment for age, sex, race, ethnicity, payer, comorbidity count, and prior utilization reduced the between-system variance from 0.566 to 0.397 and the intraclass correlation fell from 0.147 to 0.108 and the median OR from 2.05 to 1.82. Thus, two otherwise identical patients at two randomly chosen systems still differ in their odds of ever receiving a social-needs code by a median factor of about 1.8. System rankings were correspondingly stable (Spearman correlation 0.90 between crude and adjusted system ranks). With informative exceptions, rank 15 fell from 9th to 15th once its patient mix was taken into account, and rank 2 fell from 6th to 9th, while rank 13 rose from 8th to 5th. A system whose high crude rate is explained by a sicker panel with more social needs should not be penalized for coding intensity whereas a system whose coding rate remains high after adjustment is making a documentation choice, whether through screening workflows, coder training, EHR prompts, or local priority. This finding extends the system-characteristic literature,hospital Z-code use varying with organizational features,^13,14^ sparse use varying by state and domain,^12^ and rising but still selective use over time^11,15,16^ by putting an adjusted variance and an adjusted ranking on the same quantity that the outcome analyses then use.

The tertile analysis gives that variation a prognostic consequence. If high-coding systems were simply screening broadly into healthier panels, the code’s association with outcomes should dilute toward the null as coding becomes routine. For admissions, the opposite happened. Rate ratios rose from 1.20 in low-coding systems to 1.36 and 1.48 in middle- and high-coding systems (interaction P=0.0006), while ED associations did not vary materially by tertile (interaction P=0.24). At least three mechanisms could produce it, and they have different implications.

High-coding systems may document deeper into their sick panels, so that their coded population is more selected for severity even at higher coding volume. High-coding systems may also capture inpatient care more completely, through better encounter linkage or more valid stay dates, which would strengthen measured admission associations without any change in underlying risk; the episode audit argues against this potential mechanism, as discussed below, but the attenuated admission contrast in valid-date systems keeps capture differences in view.

Third, coding intensity may travel with other system practices – care management enrollment, referral completion, discharge planning – that change what happens after a code is documented. Our analyses help distinguish the first part of the question (the gradient is real and pre-specified) but not which mechanism drives it. Across the 20 matched systems, crude coding prevalence correlated only weakly with the stay-based (valid-date) share of follow-up episodes (Pearson r=0.20; Spearman ρ=0.27). This ecological check does not exclude capture differences as an explanation for the coding-intensity gradient. What should not be assumed is that raising a system’s coding rate will reproduce the risk profile of systems that already code at that rate. The newly coded patients in an expanding program are unlikely to resemble the previously coded patients, in either direction, without local evidence.

Because comparators generated 1,097 admissions per 1,000 person-years under the original counting rule – a rate that would be implausible for unselected outpatients – we audited episode construction before revising any admission claim. The audit found a record that is messy in the usual EHR way but not invalid otherwise. Only about half of EHR episodes in either analysis arm were built from valid admit and discharge dates (53.1% exposed, 46.1% comparators); the rest were calendar-day episodes. Yet only about 1% of episodes in either arm began within 1 day of the previous episode’s end, and merging episodes on that rule lowered admission rates by only about 5% in both arms, moving the adjusted admission rate ratio from 1.42 to 1.41 under the original weighting (1.43 in the corrected primary analysis) and the administrative-code ratio not at all (1.29 to 1.29). The high absolute rates may reflect the matched design doing what it was asked to do – comparators were matched to coded patients on prior ED tier, admission tier, and outpatient-volume tertile, so the comparator group is a high-utilizing outpatient population, and admitted patients in both arms averaged more than 3 admission episodes during follow-up. That interpretation is a design consequence and it is why we emphasize rate ratios and risk differences within the matched cohort over any comparison of these absolute rates to general-population benchmarks.

Medicaid patients show the strongest hardship-specific pattern (ED 1.71; admissions 1.48), which is the population where a hardship code plausibly marks the most consequential unmet need and where outreach capacity should be concentrated first. Our analyses on area-deprivation inversion – strongest associations in low-deprivation ZIP3 areas – warns against substituting an area score for the individual code, or the code for an area score. These factors measure different things, and in affluent areas a recorded code may identify a smaller, more severely selected group.

Managed-care administrators would be interested in the financial estimate of excess utilization. As gross associated acute care, the year after a code carries roughly $2,500 per patient assuming mid-range national prices, about three quarters of it preventable admissions; under an exploratory assumed-discount scenario, roughly $1,100 remains (not an estimate of hardship-attributable spending). Medicaid carries the largest ED burden. Our estimates help characterize the clinical and health care financial burden a code identifies – a caseload whose ensuing emergency and preventable hospital care is expensive – and they add support to upstream outreach and social health support programs being implemented in various settings. But these figures should not be interpreted to estimate what social health intervention may save, because the design prices prognosis, not interventions.

The code-specific gradient offers additional insights for triage and outreach programmatic activities. A homelessness code (one in six coded patients) forecasts two and a half times the ED rate and nearly twice the admission rate of matched patients – a flag that warrants same-week outreach and interventions by any plausible standard. Food insecurity is a more admission-leaning signal (its admission ratio equals its ED ratio and it feeds the preventable-admission composite), transportation and financial codes carry moderate, similar risk, and unspecified codes carry the least. Systems building work queues from Z codes should therefore sort by specific social risks documented. An opposite conclusion can also be extracted to guide program evaluation and research activities pooling Z59 into one indicator that ends up averaging distinct risks.

Health systems increasingly ask whether Z codes should enter risk prediction – as a feature in utilization models, as a stratifier for care management, or as an adjustment variable in quality comparison. Prior studies have generally shown that patients with social-needs Z codes experience higher subsequent utilization, rather than that coding improving prediction beyond clinical and utilization data. This result is consistent with the broader finding that social-needs variables in EHRs improve prediction unevenly and often modestly once utilization history is known.^34^ Our findings are compatible with both observations. A prior Z59 code identified patients with a substantially higher crude event rate, but it provided no measurable increment to a model containing demographic, comorbidity, payer, and prior-utilization variables. The discrepancy likely reflects information overlap: the circumstances that lead to documentation are correlated with prior contact, comorbidity, and acute-care history, while the code itself is rare, binary, and unevenly recorded. Our findings therefore discourage use of a single Z59 indicator as a general utilization-risk feature, while leaving open the value of richer, code-specific or longitudinal social-needs measures and the separate use of Z codes for outreach, equity monitoring, and subgroup payment assessment. A code can be clinically actionable – triggering a referral, a benefits check, a transportation voucher – without improving a discrimination metric, because actionability depends on whether an effective intervention exists for patients with social risks, not on whether the flag re-ranks the whole population.

This study extends the prior literature in at least four distinct areas. First, setting and linkage: Bensken and colleagues compared inpatients within state discharge data in 2017;^7^ Liss and colleagues estimated associations in commercial and Medicare Advantage claims;^8,9^ and the heart failure and COPD literature works within disease cohorts.^10^ The present study follows outpatients forward in linked EHR and claims records within the same 21 systems that produced the codes, so that the coding environment and the outcome environment are the same. Second, exposure specificity: most prior studies pool Z55–Z65 families. We isolate Z59 material hardship as the primary exposure, group its subfamilies (housing, transportation, food, financial, and other), and move cost-related nonadherence (Z91.120, Z91.190) to a secondary exposure on the grounds that nonadherence lies on the pathway to the outcomes. Third, the administrative-code comparison: no prior study in this literature assigns the same cohort to a hardship-free Z code.

The ratio comparing the two associations, 1.32 for ED visits, 1.11 for admissions, 1.22 for preventable admissions, 1.05 for readmission, gives readers a compact sensitivity comparison, similar to negative-control study designs; it does not isolate documentation bias.^17^ Fourth, transportability: rather than report a single prediction increment, we test whether it exists system by system, and rather than report crude coding variation, we report how much survives patient mix and how rankings move. On the other hand the null prediction increment and the valid-date admission sensitivity are both byproducts of those transportability checks, and we interpret them as part of the study’s strengths and internal consistency. Trials and evaluations of housing support, social-needs case management, and resource navigation show that acting on identified needs can change utilization in selected populations,^30–33^ and screening frameworks warn that identification without capacity to act carries its own risks.^28,29^ This study helps characterize in more detail whom the code flags and how much of the flag is documentation but it does not estimate whether acting on the flag changes outcomes. Further research is needed to answer these questions and would benefit from stepped-wedge pragmatic trial rollout designs, or carefully controlled evaluations of coding-and-referral programs that randomize or stage the intervention, not the coding.

### Strengths and Limitations

This study has several strengths structural to the analytic protocol design. The cohort rules were fixed in a versioned protocol before outcome queries, and the deviations that accumulated are listed in the Deviations from protocol v0.5 (Appendix Table S0). Matching was performed within the health system and year with a variable-ratio rule that retained 94.5% of exposed patients. Diagnosis-field matching used the enumerated raw values described in the Methods, including concatenated multi-code strings and bare parent codes. Outcomes were built from both EHR encounters and claims, deduplicated on patient-day, with an episode audit that tested the admission definition. Censoring weights were included in the primary analysis per the protocol’s 5-percentage-point rule for differential observation. Death was a screened, corroborated endpoint, and was near-identical between arms (1.54% versus 1.56% at 12 months). Finally, every primary contrast has a negative-control counterpart estimated identically, which converts the usual qualitative caveat about documentation bias into a quantitative comparison.

Among the limitations, this study estimates prognosis after documentation. It does not estimate the effect of hardship itself, or of acting on a code, and its estimates should be read as what a first Z59 code foretells in routine care. The cohort was assembled by rare-code selection, so coding prevalences describe the corpus rather than the US population; the matched contrasts are within-cohort and less exposed to that selection. Claims end in 2022 in each patient’s date frame, after which acute care is captured from health records alone; estimates from records alone were nearly identical (rate ratios 1.70 and 1.41). The 5.5% of coded patients without a comparator were heavier prior users of acute care, so our estimates are likely conservative for the full coded population. Adjustment for patient mix reduces the between-system variance by about 30% (intraclass correlation 0.147 to 0.108), which indicates that only a third of the coding spread can be attributed to case mix. The admission episode audit shows that merging episodes separated by 1 day or less lowers admission rates by about 5% in both arms and leaves the admission rate ratio essentially unchanged (1.41 under the original weighting; 1.43 in the corrected primary analysis); in the 5 systems with mostly valid stay dates the admission contrast is smaller (1.16) and no larger than its administrative-code comparison (1.29). The admission comparison was therefore attenuated in systems with better stay-date support, and the contribution of admission ascertainment to the main admission excess remains uncertain. In the unmatched yearly-index population Z code documentation added no predictive value, so the matched-cohort increment should not be used to justify risk-model adoption. Follow-up ends at the last content-bearing encounter, so patients who migrate out of a contributing system are censored by silence rather than by a disenrollment record; the censoring weights address measured predictors of that process. ZIP3 area measures use the latest recorded value and are covariates only. Systems were anonymized by rank, so we are unable to map findings to specific organizations or verify local coding initiatives. The administrative-code comparison is informative only to the extent that administrative codes share the hardship codes’ bias structure; if clinicians assign Z59 codes with different diligence than Z02/Z76 codes, the ratio comparing the two associations is an approximation, not an exact bias correction, and it does not isolate documentation bias. The primary comparator rule required no social-needs code through the end of the record; the targeted index-time sensitivity attenuated the estimates and is the appropriate check on that future-information restriction. Within the primary matched cohort no patient appears in more than one matched unit, but patient overlap between the Z59 and administrative-code cohorts could not be verified from the EMR, so the ratio intervals, which treat the two cohorts as independent, carry that unverified assumption. Claims-based inpatient dates are observable only through 2022 and are merged with EHR stays before any 365-day outcome window is applied, so no outcome depends on claims capture beyond the record; the EHR-only sensitivity bounds any residual capture difference. Finally, the corpus ends claims capture in 2022 and shifts every patient’s dates, so results can not be interpreted under the context of a policy change, or a pandemic period. The cost analysis also inherits the previous data limitations. A payer is claim-header text, preventable-admission dollars by payer are allocations, and prices are national planning values rather than payments observed in this corpus

### Conclusions

Systems assign hardship codes at rates that differ more than twentyfold for reasons that patient mix only partly explains. A first material-hardship code flags patients who will use substantially more emergency and hospital care over the next year, in every system studied. A paperwork-only code flags some of the same risk for admissions but no hardship-specific excess was detectable for admissions where stay dates were most complete. Health systems should therefore treat a Z59 code as a strong local risk signal but interpret it based on its specific coding culture. Coding is instrumental for identifying patients for outreach and social health programming. Raw coding rates should not be used to compare systems, Z59 indicators should not be expected to improve a risk model that already knows prior utilization, and every admission association should be read alongside its administrative-code comparison.

## Policy recommendations

(1) Do not benchmark raw coded rates across systems (23-fold range; median odds ratio 1.82 after patient mix). (2) Triage outreach and intervention plans by which social-need codes were recorded: homelessness first (ED 2.49), then inadequate housing, food, transportation, and financial hardship. (3) Do not add a Z59 indicator to a utilization risk score that already includes prior utilization (4) Fund Medicaid outreach first (strongest signal, ED 1.71 and admissions 1.48; largest ED burden, 954 per 1,000 person-years). (5) Before Z codes enter quality measurement or risk adjustment engines, define what a code certifies: the same flag means different things under different coding cultures, and every admission association needs its administrative-code comparison.

## Acknowledgements

The authors thank the Meta Superintelligence Labs for data access and technical support.

## Use of Artificial Intelligence

This study was supported through an internal Meta scaffold around Muse Spark 1.3 and Wonder, which assisted the authors in all stages of the study. The models were used to draft the analyses, study cohorts, statistical plan, protocols, and manuscript preparation. The authors drafted all inputs to the models, and all output was verified by the lead author. Analyses followed a time-stamped versioned protocol (v0.5); deviations are listed in Appendix Table S0, and analyses not prescribed in the protocol are denoted exploratory.

## Author Contributions

F.L. conceived and designed the study, supervised the analyses, interpreted the data, and wrote the manuscript. J.D. contributed to the statistical analysis plan and data interpretation. S.A. provided critical review of the manuscript and interpretation. C.C. provided study coordination, data access and critical interpretation and manuscript review. All authors reviewed and edited the manuscript and approved the final version. F.L had full access to all the data in the study through the governed analysis environment.

## Data Availability

The Protégé data are not publicly available and access is governed by the data owner. The study protocol, statistical analysis plan, code lists and aggregate results are available from the Corresponding author on reasonable request.

## STROBE Cohort and RECORD Checklist — Social Needs Z Codes and Subsequent Acute Care Across 21 U.S. Health Systems

| **Item** | **Recommendation** | **Status** | **Where reported** |
| --- | --- | --- | --- |
| STROBE 1a<br>Title/abstract | Indicate study design in title or abstract | Addressed | Title; Abstract (matched cohort, negative control) |
| STROBE 1b Abstract | Balanced summary of what was done and found | Addressed | Abstract |
| STROBE 2<br>Background | Scientific background and rationale | Addressed | Introduction |
| STROBE 3<br>Objectives | Specific objectives, including hypotheses | Addressed | Introduction, final paragraphs |
| STROBE 4 Design | Key elements of study design early | Addressed | Methods, Study Sample |
| STROBE 5 Setting | Setting, locations, relevant dates | Addressed with data limit | Methods (21 data owners, shifted years 2013-2026, snapshot 2026-06-04; no real-calendar date) |
| STROBE 6a<br>Participants | Eligibility, sources, selection | Addressed | Methods; Supplement eTable 3 |
| STROBE 6b Matched studies | Matching criteria and numbers | Addressed | Methods; Table 1 (98,869 comparator units, weighted 26,153) |
| STROBE 7 Variables | Outcomes, exposures, predictors, confounders, effect modifiers | Addressed | Measures; Supplementary Code List eTable 1 |
| STROBE 8<br>Measurement | Data sources and measurement for each variable | Addressed | Measures; eTables 1-3 |
| STROBE 9 Bias | Efforts to address potential bias | Addressed | Negative control; IPCW; episode audit; Discussion |
| STROBE 10 Study size | How study size was arrived at | Addressed | Cohort flow in Methods and Results |
| STROBE 11 Quantitative variables | Groupings and handling | Addressed | Measures; Statistical Analysis |
| STROBE 12a-e Statistical methods | Methods, confounding, subgroups, missing data, sensitivity | Addressed | Statistical Analysis; Results sensitivities |
| STROBE 13a-c Participants | Numbers at each stage, flow, follow-up | Addressed | Results; Table 1 unmatched column |
| STROBE 14a-c Descriptive data | Characteristics, missing data, follow-up time | Addressed | Table 1; Methods (observation) |
| STROBE 15 Outcome data | Outcome events over time | Addressed | Results; Table 2 |
| STROBE 16a-c Main results | Unadjusted/adjusted estimates, precision | Addressed | Results; Tables 2-3 |
| STROBE 17 Other analyses | Subgroup, interaction, sensitivity | Addressed | Results; Supplement S3-S7 |
| STROBE 18 Key results | Summary referencing objectives | Addressed | Discussion opening |
| STROBE 19 Limitations | Limitations, bias direction/magnitude | Addressed | Limitations |
| STROBE 20 Interpretation | Cautious overall interpretation | Addressed | Discussion; Conclusions |
| STROBE 21 Generalisability | External validity | Addressed | Limitations (enrichment; prevalences corpus-only) |
| STROBE 22 Funding | Source of funding and role | Addressed | Acknowledgments/Funding (author completion) |
| RECORD 6.1 | Codes and algorithms to identify population, exposures, outcomes, covariates | Addressed | Supplementary Code List eTable 1 (433 rows) |
| RECORD 7.1 | List of codes and definitions, with validation where available | Addressed with gaps marked | eTables 1-3; |
| RECORD data linkage | Linkage methods and quality | Addressed | Methods (EHR-claims linkage, patient-day deduplication) |
| Protocol deviations | Pre-registration and deviations | Deviation, transparently reported | Appendix Table S0 (Deviations from protocol v0.5, below) |

## Appendix methods

Censoring model, weight capping at the 99.5th percentile, 500 matched-set bootstrap, episode construction and audit (stay-based share 53.1% exposed vs 46.1% comparators; within-1-day starts about 1%), matched-cohort prediction sensitivity (ΔAUC 0.0074, sensitivity only), and the coding-capture mechanism check (prevalence vs stay-based share Pearson r=0.20, Spearman 0.27; tertile weighted stay share 0.36/0.61/0.46) are described in the deposited reports and code.

## Deviations from protocol v0.5

1. Prediction analysis initially used the matched cohort instead of the unmatched yearly-index population; corrected, protocol-population increment is null (ΔAUC 0.0000).
2. Leave-one-system-out validation was initially dropped over sparse demographic levels instead of collapsing categories under 1%; corrected (ΔAUC -0.0003 to 0.0002 by system).
3. Between-system variance was initially reported unadjusted for patient mix; corrected (intraclass correlation 0.11 adjusted).
4. Secondary outcomes (Prevention Quality Indicators and 30-day readmission) now reported.
5. A targeted index-time comparator-eligibility sensitivity was added after review (Supplement Table S3); the primary comparator rule (no family code through end of record) is unchanged in the primary analysis.
6. Site-variance headline initially used a prevalence meta-analysis instead of the specified multilevel logistic model

**Appendix Figure 1.**
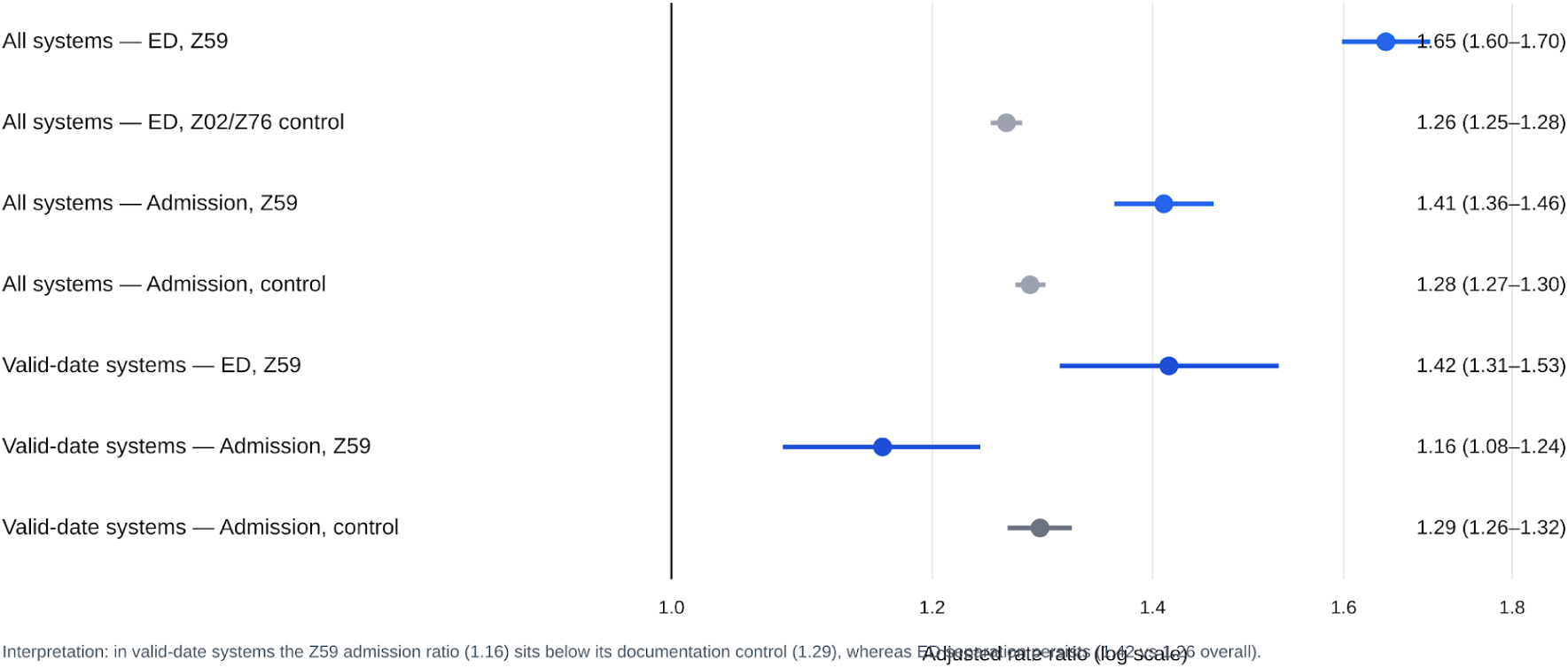
Valid-date sensitivity: in the 5 systems where at least half of inpatient rows carry valid stay dates, the hardship admission ratio (1.16) sits below its paperwork control (1.29) while emergency separation persists;

## Supplement Tables S1-S7

**Supplement Table S1.** Code lists (the Supplementary Code List, eTables 1-3, is a separate submission file and is not reproduced in this combined document)

| Component | Codes / definition | Source |
| --- | --- | --- |
| Primary exposure | 38 Z59 material-hardship codes (Z59 family incl. bare Z59) | zcodes_normalized.csv |
| Secondary exposure | Z91.120, Z91.190 (cost-related nonadherence) | — |
| Negative control | Z02/Z76 administrative codes | negctrl_normalized.csv |
| Preventable admissions | AHRQ Prevention Quality Indicators<br>ICD-10-CM/PCS v2025 composite (PQI 01, 03, 05, 07, 08, 11, 12, 14, 15) | pqi_acsc_codes.csv |
| PQI sensitivity | PQI 10 dehydration v7.0 | pqi10_codes.csv |
| Comorbidities | Elixhauser comorbidities (Quan et al. prefix algorithms) | — |

**Supplement Table S2.** Unmatched exposed patients and match retention.

| Metric | Value |
| --- | --- |
| Eligible exposed patients | 27,683 |
| Retained at least one comparator | 26,153 (94.5%) |
| Remained unmatched | 1,530 (5.5%) |
| Comparator units | 98,869 |
| Weighted comparator total | 26,153 |
| Matching | Variable-ratio, up to 1:4; unmatched patients were heavier prior users (see main Table 1, unmatched column) |

**Supplement Table S3.** Sensitivity analyses (rate ratios, Z59 vs comparators) Note: Corrected primary estimates use rebuilt recurrent-event censoring weights (bootstrap intervals). Consistent event/person-time (Poisson) sensitivity: ED 1.60 (1.54-1.75), admissions 1.26 (1.18-1.35); the lower admission figure mainly reflects the model change, not the weight correction. Overlap audit: the primary matched cohort contains 125,022 units and 125,022 unique patients (no repeated comparator units, no dual-role patients); in the index-time sensitivity cohort 4,714 patients appear in both roles (comparator before their own first Z59 code), 3,513 with overlapping outcome windows. Patient overlap between the Z59 and administrative-code cohorts could not be counted because the administrative patient-level match files are not in the deposited record; the joint patient-cluster bootstrap was therefore not run and the independent-cohort ratio intervals remain unverified. Targeted index-time sensitivity (26,694 exposed patients retained; 101,546 comparator units; same pipeline): ED 1.55 (1.50-1.60), admissions 1.33 (1.28-1.37), against 1.65 and 1.42 under the original through-end-of-record eligibility in the same pipeline; first-event risk difference 11.7 vs 13.1 percentage points.

| Analysis | ED rate ratio | Admission rate ratio | Notes |
| --- | --- | --- | --- |
| Unweighted | 1.66 | 1.42 |  |
| EHR-only | 1.70 | 1.41 |  |
| Complete 1:4 sets | 1.68 | 1.42 |  |
| 1:2 sets | 1.66 | 1.41 |  |
| Excluding 2 lowest-retention systems | 1.66 | 1.42 |  |
| Adding 5 imbalanced covariates | 1.61 | 1.41 |  |
| Valid-date systems (5) | — | 1.16 | Administrative-code comparison 1.29 (admission ascertainment uncertain) |
| Primary (merged episodes) | 1.67 (1.61-1.74) | 1.43 (1.38-1.49) |  |

**Supplement Table S4.** Prediction performance (protocol population, 298,070 patient-years)

| Metric | Result | Interpretation |
| --- | --- | --- |
| Baseline model AUC (protocol population) | 0.7594 | 298,070 patient-years |
| Change in AUC after adding prior-12-month Z59 | 0.0000 | No increment |
| Integrated discrimination improvement | 0.000008 |  |
| Categorical NRI at 5%, 10%, 20% thresholds | ~0 |  |
| Leave-one-system-out change in AUC | -0.0003 to 0.0002 | All systems |
| Matched-cohort sensitivity: baseline AUC | 0.7414 | No increment |

**Supplement Table S5.**
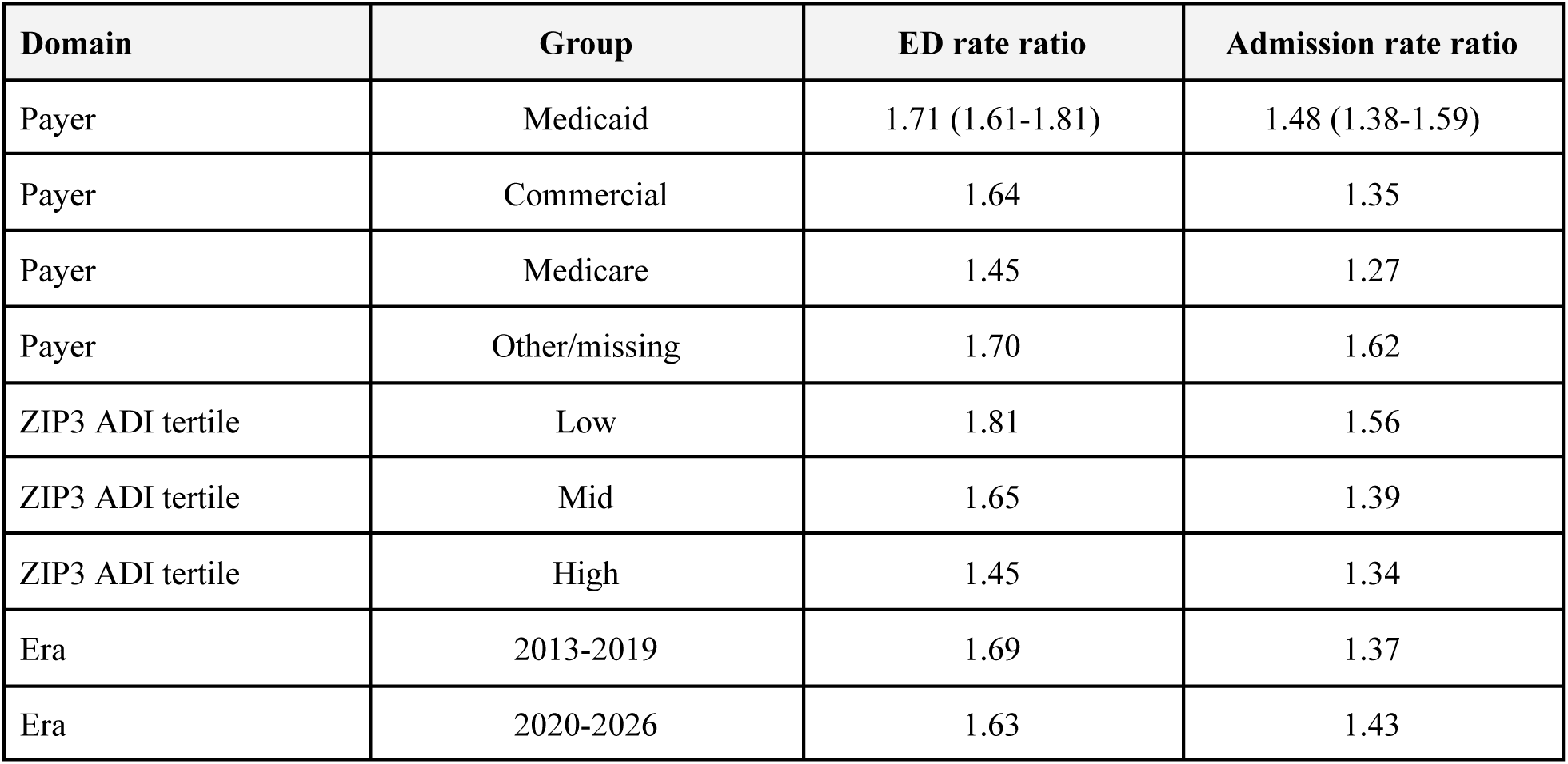
Stratified sensitivities.

| Domain | Group | ED rate ratio | Admission rate ratio |
| --- | --- | --- | --- |
| Payer | Medicaid | 1.71 (1.61-1.81) | 1.48 (1.38-1.59) |
| Payer | Commercial | 1.64 | 1.35 |
| Payer | Medicare | 1.45 | 1.27 |
| Payer | Other/missing | 1.70 | 1.62 |
| ZIP3 ADI tertile | Low | 1.81 | 1.56 |
| ZIP3 ADI tertile | Mid | 1.65 | 1.39 |
| ZIP3 ADI tertile | High | 1.45 | 1.34 |
| Era | 2013-2019 | 1.69 | 1.37 |
| Era | 2020-2026 | 1.63 | 1.43 |

**Supplement Table S6.** Administrative-code comparison (observed-comparison shares; not identified documentation effects)

| Outcome | Ratio comparing the two associations (95% CI) | Observed-comparison share of Z59 log-association |
| --- | --- | --- |
| ED visits | 1.32 (1.27-1.38) | 46% |
| Admissions (merged) | 1.11 (1.07-1.16) | 70% |
| PQI composite | 1.22 (1.15-1.30) | 61% |
| PQI 10 | 1.26 (1.13-1.40) | 61% |
| Readmission episodes | 1.05 (0.97-1.14) | 87% |
| Any readmission among admitted (OR) | 0.96 (0.93-0.99) | >100% |

**Supplement Table S7.** Modeled excess costs per coded patient-year (2023 USD)

| Component | Excess events /1,000 py | Gross costs per coded patient-year | Assumed-discount scenario costs (exploratory; not hardship-attributable estimates) |
| --- | --- | --- | --- |
| Emergency department visits | 654 | \$654 | \$355 |
| Preventable admissions | 124 | \$1,862 | \$719 |
| Total, base prices (\$1,000 ED; \$15,000 stay) | — | \$2,516 | \$1,074 |
| Total, low prices (\$600; \$11,000) | — | \$1,758 | \$739 |
| Total, high prices (\$1,600; \$20,000) | — | \$3,529 | \$1,525 |

**By payer (base prices, payer-adjusted). This panel shows the ED component only; payer-specific preventable-admission dollars are modeled allocations in the deposited cost_by_payer.csv and are not measured cells in this table.**

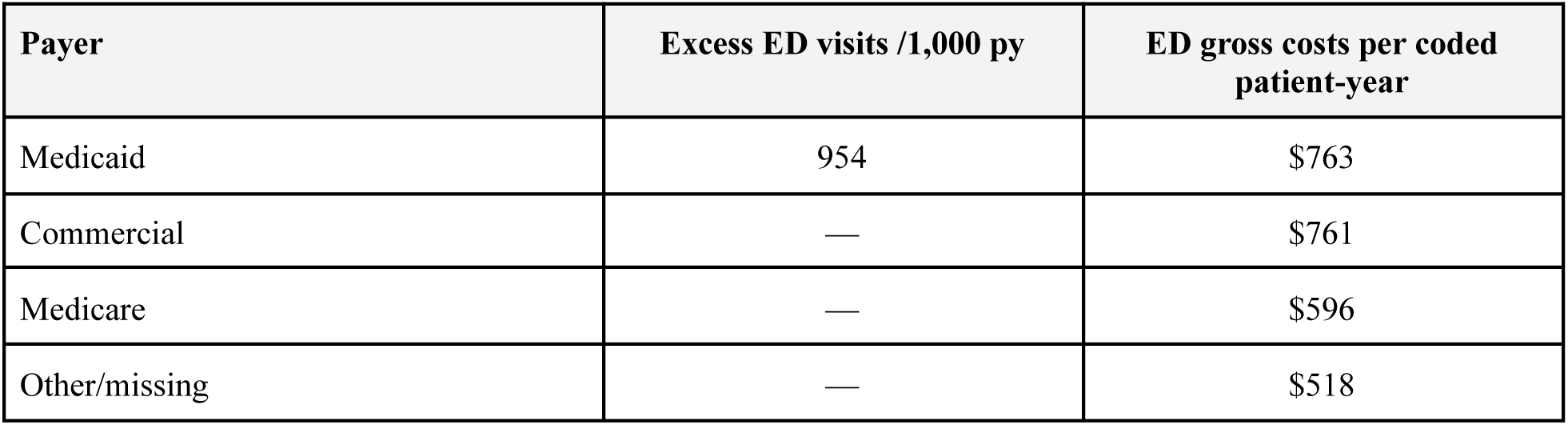

## References

1. Braveman P, Gottlieb L. The social determinants of health: it’s time to consider the causes of the causes. Public Health Rep. 2014;129(suppl 2):19–31. doi:10.1177/00333549141291S206

2. Solar O, Irwin A. A Conceptual Framework for Action on the Social Determinants of Health. Geneva, Switzerland: World Health Organization; 2010.

3. National Academies of Sciences, Engineering, and Medicine. Integrating Social Care into the Delivery of Health Care: Moving Upstream to Improve the Nation’s Health. Washington, DC: The National Academies Press; 2019.

4. Gottlieb LM, Wing H, Adler NE. A systematic review of interventions on patients’ social and economic needs. Am J Prev Med. 2017;53(6):719–729. doi:10.1016/j.amepre.2017.07.009

5. Torres JM, Lawlor J, Colvin JD, et al. ICD social codes: an underutilized resource for tracking social needs. Med Care. 2017;55(9):810–816. doi:10.1097/MLR.0000000000000764

6. Centers for Medicare & Medicaid Services, Office of Minority Health. Using Z Codes: The Social Determinants of Health (SDOH) Data Journey to Better Outcomes. Baltimore, MD: Centers for Medicare & Medicaid Services; 2023.

7. Bensken WP, Alberti PM, Stange KC, et al. ICD-10 Z-code health-related social needs and increased healthcare utilization. Am J Prev Med. 2022;62(4):e232–e241. doi:10.1016/j.amepre.2021.10.004

8. Liss DT, Kang RH, Cherupally M, et al. Association between ICD-10 codes for social needs and subsequent emergency and inpatient use. Med Care. 2024;62(1):60–66. doi:10.1097/MLR.0000000000001948

9. Liss DT, Cherupally M, Kang RH, et al. Social needs identified by diagnostic codes in privately insured U.S. adults. Am J Prev Med. 2022;63(6):1007–1016. doi:10.1016/j.amepre.2022.07.009

10. Z-code health-related social needs and their association with post-discharge healthcare utilization in older adults with heart failure and COPD. J Gen Intern Med. 2025. doi:10.1007/s11606-025-09887-8 [Full author list to verify against publisher record before submission.]

11. Bensken WP, Alberti PM, Baker MC, Koroukian SM. An increase in the use of ICD-10 Z-codes for social risks and social needs: 2015 to 2019. Popul Health Manag. 2023;26(2):89–97. doi:10.1089/pop.2022.0248

12. Aswani MS, Do LA, Shafer PR. Use of social determinants of health Z codes was sparse, 2016-22. Health Aff (Millwood). 2025;44(5):631–635. doi:10.1377/hlthaff.2024.01033

13. Chang JE, Smith N, Lindenfeld Z, Weeks WB. Hospital use of common Z-codes for Medicare fee-for-service beneficiaries, 2017-2021. Health Aff Sch. 2024;2(1):qxad086. doi:10.1093/haschl/qxad086

14. Utilization of social determinants Z codes and health system characteristics, 2015-2024. Am J Manag Care. 2025-2026. [Full citation and author list to confirm against the current AJMC record before submission.]

15. Weeks WB, Cao SY, Lester CM, et al. Use of Z-codes to record social determinants of health among fee-for-service Medicare beneficiaries in 2017. J Gen Intern Med. 2020;35(3):952–955. doi:10.1007/s11606-019-05502-w

16. Truong HP, Luke AA, Hammond G, et al. Utilization of social determinants of health ICD-10 Z-codes among hospitalized patients in the United States, 2016-2017. Med Care. 2020;58(12):1037–1043. doi:10.1097/MLR.0000000000001410

17. Lipsitch M, Tchetgen Tchetgen E, Cohen T. Negative controls: a tool for detecting confounding and bias in observational studies. Epidemiology. 2010;21(3):383–388. doi:10.1097/EDE.0b013e3181d61eeb

18. Quan H, Sundararajan V, Halfon P, et al. Coding algorithms for defining comorbidities in ICD-9-CM and ICD-10 administrative data. Med Care. 2005;43(11):1130–1139. doi:10.1097/01.mlr.0000182534.19832.83

19. Agency for Healthcare Research and Quality. Prevention Quality Indicators Technical Specifications, ICD-10-CM/PCS v2025. Rockville, MD: Agency for Healthcare Research and Quality; 2025.

20. Cole SR, Hernán MA. Constructing inverse probability weights for marginal structural models. Am J Epidemiol. 2008;168(6):656–664. doi:10.1093/aje/kwn164

21. Aalen OO, Johansen S. An empirical transition matrix for non-homogeneous Markov chains based on censored observations. Scand J Statist. 1978;5(3):141–150.

22. VanderWeele TJ, Ding P. Sensitivity analysis in observational research: introducing the E-value. Ann Intern Med. 2017;167(4):268–274. doi:10.7326/M16-2607

23. Merlo J, Chaix B, Ohlsson H, et al. A brief conceptual tutorial of multilevel analysis in social epidemiology: using measures of clustering in multilevel logistic regression to investigate contextual phenomena. J Epidemiol Community Health. 2006;60(4):290–297. doi:10.1136/jech.2005.040253

24. Pencina MJ, D’Agostino RB Sr, D’Agostino RB Jr, Vasan RS. Evaluating the added predictive ability of a new marker: from area under the ROC curve to reclassification and beyond. Stat Med. 2008;27(2):157–172. doi:10.1002/sim.2988

25. Vickers AJ, Elkin EB. Decision curve analysis: a novel method for evaluating prediction models. Med Decis Making. 2006;26(6):565–574. doi:10.1177/0272989X06295361

26. von Elm E, Altman DG, Egger M, et al. The Strengthening the Reporting of Observational Studies in Epidemiology (STROBE) statement: guidelines for reporting observational studies. Lancet. 2007;370(9596):1453–1457. doi:10.1016/S0140-6736(07)61602-X

27. Benchimol EI, Smeeth L, Guttmann A, et al. The REporting of studies Conducted using Observational Routinely-collected health Data (RECORD) statement. PLoS Med. 2015;12(10):e1001885. doi:10.1371/journal.pmed.1001885

28. Moons KGM, Altman DG, Reitsma JB, et al. Transparent Reporting of a multivariable prediction model for Individual Prognosis or Diagnosis (TRIPOD): explanation and elaboration. Ann Intern Med. 2015;162(1):W1–W73. doi:10.7326/M14-0698

29. Garg A, Boynton-Jarrett R, Dworkin PH. Avoiding the unintended consequences of screening for social determinants of health. JAMA. 2016;316(8):813–814. doi:10.1001/jama.2016.9282

30. Andermann A. Taking action on the social determinants of health in clinical practice: a framework for health professionals. CMAJ. 2016;188(17-18):E474–E483. doi:10.1503/cmaj.160177

31. Parish W, Beil H, He F, et al. Health care impacts of resource navigation for health-related social needs in the Accountable Health Communities model. Health Aff (Millwood). 2023;42(6):822–831. doi:10.1377/hlthaff.2022.01502

32. Brown DM, Hernandez EA, Levin S, et al. Effect of social needs case management on hospital use among adult Medicaid beneficiaries. Ann Intern Med. 2022;175(8):1109–1117. doi:10.7326/M22-0078

33. Sadowski LS, Kee RA, VanderWeele TJ, et al. Effect of a housing and case management program for emergency department visits and hospitalizations among chronically ill homeless adults: a randomized trial. JAMA. 2009;301(17):1771–1778. doi:10.1001/jama.2009.561

34. Hollander MAG, Cole ES, Donohue JM, et al. Changes in Medicaid utilization and spending associated with homeless adults’ entry into permanent supportive housing. J Gen Intern Med. 2021;36(8):2353–2360. doi:10.1007/s11606-020-06524-4

35. Chen M, Tan X, Padman R. Social determinants of health in electronic health records and their impact on analysis and risk prediction: a systematic review. J Am Med Inform Assoc. 2020;27(11):1764–1773. doi:10.1093/jamia/ocaa109

36. Steyerberg EW, Vickers AJ, Cook NR, et al. Assessing the performance of prediction models: a framework for traditional and novel measures. Epidemiology. 2010;21(1):128–138. doi:10.1097/EDE.0b013e3181c30fb2

37. Healthcare Cost and Utilization Project (HCUP). Costs of Treat-and-Release Emergency Department Visits in the United States, 2021. Statistical Brief No. 311. Rockville, MD: Agency for Healthcare Research and Quality; 2024.

38. Agency for Healthcare Research and Quality. HCUP National Inpatient Sample: average costs of inpatient stays. Rockville, MD: Agency for Healthcare Research and Quality; 2023-2024 releases. [Confirm exact release at submission.]

39. Health Care Cost Institute. Health Care Cost and Utilization Report. Washington, DC: Health Care Cost Institute; 2024. [Confirm edition at submission.]

